# The impact of mandatory COVID-19 certificates on vaccine uptake: Synthetic Control Modelling of Six Countries

**DOI:** 10.1101/2021.10.08.21264718

**Authors:** Melinda C. Mills, Tobias Rüttenauer

## Abstract

**Background:** COVID certification has been introduced, yet there are no empirical evaluations of its impact on vaccine uptake.

**Methods:** Mirroring an RCT, we designed a synthetic control model comparing six countries (Denmark, Israel, Italy, France, Germany, Switzerland) that introduced certification (May-August 2021), with 20 control countries. Our estimates provide a counterfactual trend estimating what would have happened in virtually identical circumstances if certificates were not introduced. The primary outcome was daily COVID-19 vaccine doses, with supplementary analyses of COVID-19 infections.

**Findings:** COVID-19 certification led to increased vaccinations 20 days prior to implementation, with a lasting effect up to 40 days after. Countries with lower than average pre-intervention uptake had a more pronounced increase. In France, doses exceeded 25,895 vaccines per million capita (pmc) or in absolute terms, 1,749,589 doses prior to certification and 11,434 pmc after (772,563 doses). There was no effect in countries with higher uptake (Germany) or when introduced during limited supply (Denmark). There was higher uptake for <20 years and 20-29 years. Access restrictions linked to certain settings (nightclubs, events >1,000) were associated with higher uptake <20 years. When extended to broader settings, uptake remained high in the youngest group, but also observed in older age groups. The relationship of the intervention with reported infections was difficult to assess based on available data.

**Interpretation:** We provide the first empirical assessment of the relationship between COVID-19 certification and vaccine uptake. Interpretation should recognise additional factors, including age eligibility changes and pandemic trajectories. We provide evidence that certification could increase vaccine uptake.

**Funding and Competing Interest Statement:** MCM receives funding from the Leverhulme Trust (Large Centre Grant), European Research Council (835079) and participates in UK’s SAGE SPI-B (behavioural insights) committee. The funders had no role in study design, data collection, analysis, interpretation, or writing of the report.

**Research in Context:** *Evidence before this study:* The introduction of COVID-19 certification or vaccine passports has been linked to lower self-reported vaccine intentions, yet national media and health offices report increases in vaccinations. No empirical studies could be located that had examined the impact of the implementation of mandatory COVID-19 certification on vaccine uptake.

*Added value of this study:* To our knowledge, this is the first empirical analysis of the relationship of the introduction of COVID-19 certification on vaccine uptake.

*Implications of all the available evidence:* Our study provides the first evidence that mandatory COVID-19 certification restricting access to certain settings can influence vaccine uptake for those groups affected by the intervention. Given higher vaccine complacency in certain groups, such as youth who perceive lower risks of infection, this intervention could be an additional policy lever to increase vaccine uptake and population level immunity. Future studies examining more countries and variation by eligibility criteria and factors beyond age are warranted.

## Introduction

Countries have either introduced or are considering mandatory COVID-19 certification using proof of a double vaccination, negative PCR or lateral flow test, or a viral antibody serological test to demonstrate recent natural infection.^1,2^ Certification manages entry into settings such as nightclubs, large events, hospitals, gyms or indoor hospitality (see Supplementary Material, Table A1). Numerous media reports have linked certification to increased vaccine uptake,^3–5^ yet evidence remains anecdotal. Self-reported surveys suggest certification lowers vaccine intentions,^6^ and a recent systematic review,^7^ concluded that the quality and quantity of studies was low and concluded there was no evidence it would increase vaccine uptake.^7^

Given that certain groups, such as youth, men and some ethnic minorities have lower levels of vaccine uptake – often attributed to vaccine complacency and lower perceptions of risk from COVID-19^8,9^ – a relevant question is whether certification could be an additional policy lever to increase uptake. Vaccines provide not only protection for the immunised, but reduces transmission and risk of serious illness and death and only when sufficient numbers are vaccinated, can the chain of infection be broken.^10^ Given mixed findings and lack of empirical evidence, we conducted the first empirical study to assess the relationship between the introduction of COVID-19 certification on observed vaccine uptake from May to 29 August 2021.

## Methods

### Study design and data

Information on COVID-19 related health indicators was used from Our World in Data,^11,12^ which harmonises daily information on indicators such as cases, deaths, and vaccinations. We linked these to the Oxford COVID-19 Government Response Tracker (OxCGRT),^13^ which provides daily country information on the implementation and stringency of non-pharmaceutical interventions. Age-specific analyses used European Centre for Disease Prevention and Control (ECDC) data^14^ on weekly age-specific vaccine doses. For all sources, we used the data around the time of COVID certification in each country in 2021 until 28 September 2021.

### COVID-19 certification

We selected six cases of Denmark, France, Germany, Israel, Italy and Switzerland, that implemented this intervention between May and August 2021, where sufficient data and time was available to examine effects. Details on when each intervention was announced, introduced, setting, rules or exemptions is in Supplementary Material Table A1, with the timing shown in Figure 1.

**Figure 1.**
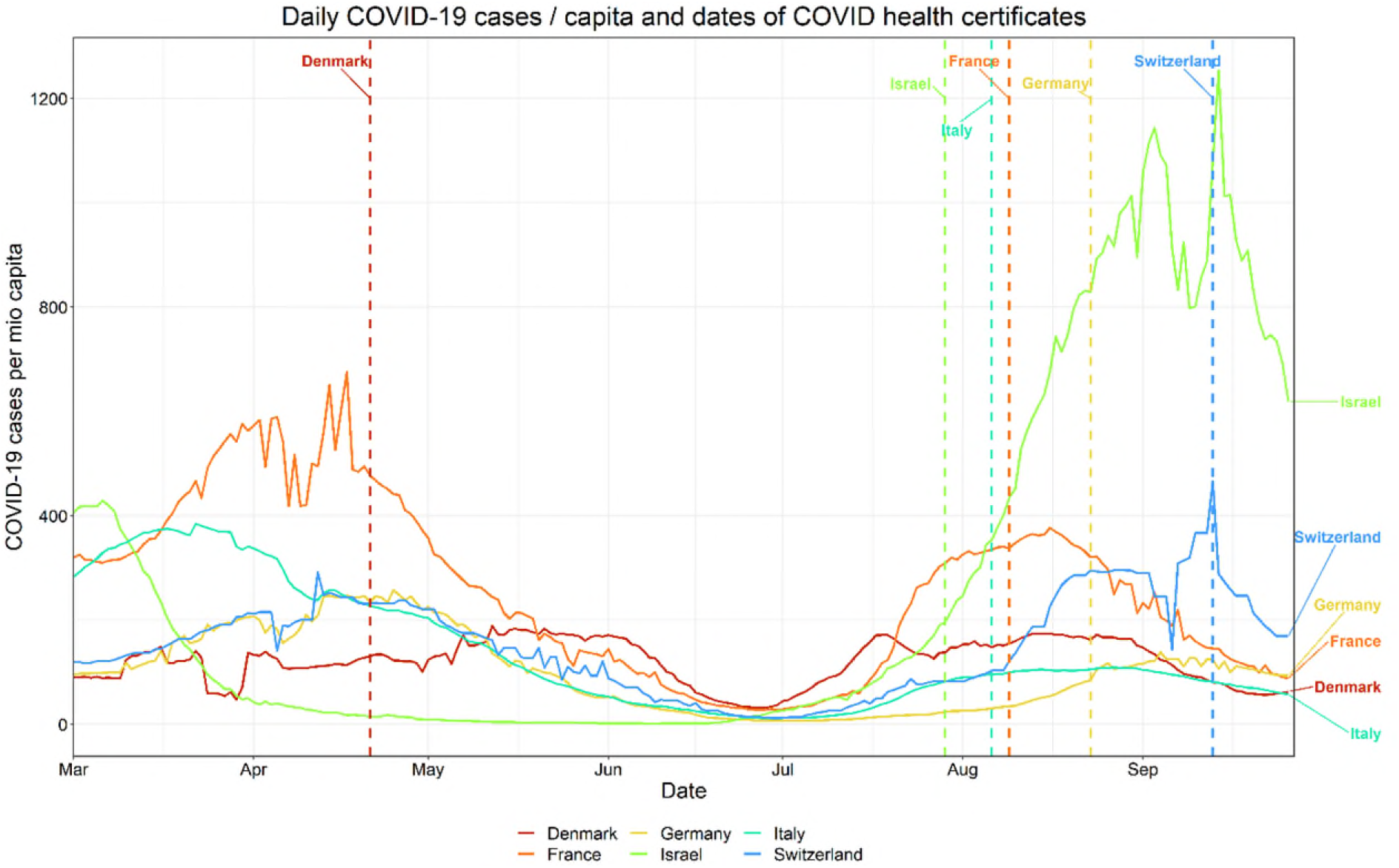
COVID-19 cases by selected countries that introduced COVID certification showing date of introduction (vertical line), March to 28 September 2021 Note: Implementation dates are: Denmark: 2021-04-21 (most restrictions lifted in August/September 2021); Israel: 2021-07-29 (re-introduction of the certificate); Italy: 2021-08-06; France: 2021-08-09; Germany: 2021-08-23; Switzerland: 2021-09-13.

A country was considered as implementing mandatory certificates if access to at least some frequently used public venues such as restaurants, or cultural events was restricted to people with a certificate (see Table A1). Some countries brought in certificates earlier, but did not associate restrictions or only applied rules to specific settings (e.g. Switzerland, larger events since 01 July). In the main analysis, we focus only on general access restrictions and examine different settings in a secondary analysis. In some cases, certificates were introduced gradually in different settings such as in Denmark in some settings on 6 April (e.g., hairdressers) when most public places were closed and later extended to most public places (21 April).

### Outcomes

The primary outcome is daily new COVID-19 vaccination doses administered per capita, using a 7-day smoothed rolling average.^12^ For age-specific vaccination doses, we allocated the age-specific weekly doses (by ECDC)^14^ to each day of the week proportionate to each day’s share of total doses administered in the respective week (by Our World in Data). For instance, if there were 1,000 doses amongst the age group 18-24 in week 30 (ECDC), and 25% of all vaccine doses in this week were given on a Wednesday (Our World in Data), we allocated 250 doses for the age group 18-24 to Wednesday, and so forth. In secondary analyses, we tested the effect of certificates on daily reported infection COVID cases. All health-related outcomes were adjusted by (age-specific) population (per million). Harmonized age-specific vaccination rates from the ECDC were only available for France, Italy, and Denmark among the treated countries.

Age-specific vaccination data from the ECDC provides harmonized data, allowing us to run Synthetic Control models (see below) on age-specific vaccination rates. However, data is only available for relatively coarse age groups. We therefore compiled original data on detailed age-specific vaccination rates directly from national sources for France,^15^ Israel,^16^ Italy,^17^ and Switzerland.^18^ Since data from Israel came without age-specific counts of the total population, we supplemented this population information.^19^ For Switzerland, the age-specific vaccination data is only available on a weekly basis. We therefore applied the same method as described above for the ECDC data.

### Statistical analyses

We used a Synthetic Control method, which constructs a synthetic control group for each single treated country^20,21^ by reweighting non-treated countries from the potential pool of control units in a way that the average pre-treatment trend (and other selected characteristics) closely mirroring the trend of the single treated unit. The synthetic control group average provides a counterfactual trend of the outcome for the treated unit. We estimate the effect of certificates on new daily vaccine doses.

Matching was performed on time-constant country characteristics: (1) median age, (2) percent 70 and older, (3) life expectancy, (4) GDP per capita; and, (5) population density and time-varying characteristics: (6) pre-treatment outcome (8-14, 15-21, 22-28 days before intervention); (7) stringency sub-indices of 12 NPIs (8-21 days before intervention), including: school closing, workplace closing, cancelled public events, restricted gatherings, closed public transport, stay at home, restricted internal movement, international travel, protecting elderly, testing policy, contact tracing, face coverings; (8) daily COVID-19 cases (1-7, 8-14, 15-21, 22-28 days before intervention); and, (9) percent fully vaccinated (8-21 days before intervention).

In a secondary analyses we examine COVID-19 reported infection cases as the outcome, including the following variables (instead of daily cases and percent fully vaccinated): (1) cumulative vaccinations per 100 inhabitants (1-7, 8-14, 15-21, 22-28 days before intervention); and, (2) daily tests per capita performed (8-21 days before intervention).

The following countries that did not introduce certification are included in the pool of control countries: Austria, Belgium, Canada, Czech Republic, Spain, Finland, Great Britain, Turkey, Croatia, Ireland, Lithuania, Luxembourg, Netherlands, Norway, Poland, Portugal, Slovakia, Slovenia, Sweden, and the United States. Due to data availability in some control countries, the following countries were included as control units for the age-specific analyses: Austria, Belgium, Cyprus, Czech Republic, Spain, Estonia, Finland, Turkey, Croatia, Ireland, Iceland, Lithuania, Luxembourg, Latvia, Malta, Norway, Poland, Portugal, Romania, Slovakia, Slovenia, and Sweden. We note that the results look similar for these two sets of control units. Analyses were performed using the R package gsynth,^22^ and results visualized using ggplot.^23^

In supplementary analyses, we used the Generalized Synthetic Control (GSC) method^24^ to estimate an average effect across our selected cases in comparison with remaining countries included in Our World in Data. The GSC method uses the information of control units, and the pre-treatment trends of the treated units to impute the expected outcome of the treated units after receiving the intervention.^25^ As covariates, we included the predictors mentioned above on a daily basis and two-ways fixed effects to account for country-specific differences and time-specific shocks.

## Results

We first assess the relationship between COVID-19 certification and overall vaccine uptake per capita. Figure 2 shows development over time of new daily vaccinations per million by the treated country (with certification) and synthetic control group (countries with no certification) (left panel) in France. The right panel illustrates difference in daily vaccine doses per million around the day of mandatory certification (*pass sanitaire*) 09 August 2021. France had a lower rate of daily vaccinations per capita compared to other similar countries until three weeks prior to the intervention. When mandatory certificates were announced in mid-July, daily vaccinations started to rise to an above-average level and remained there after certificates were required.

**Figure 2.**
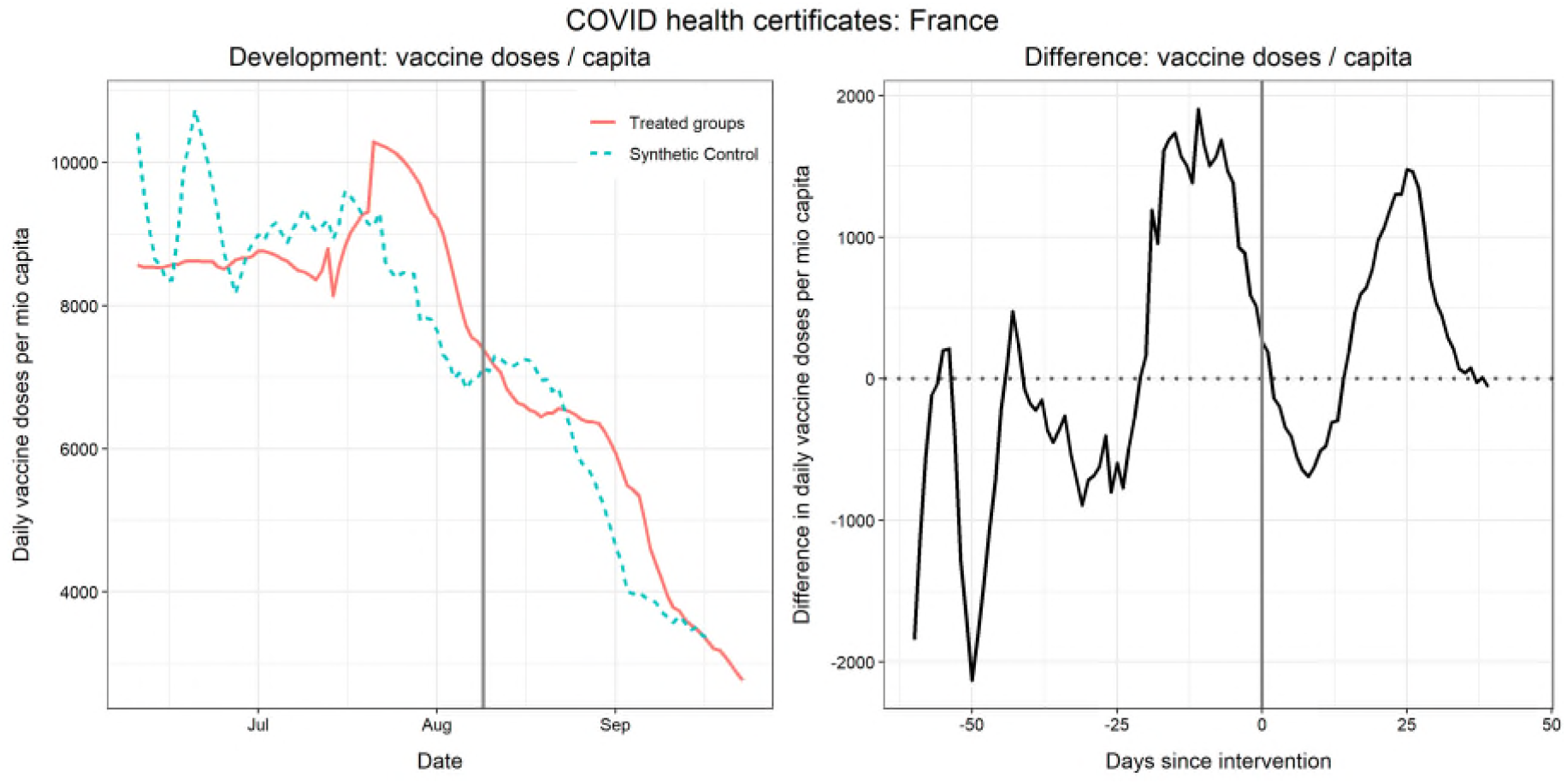
Daily new vaccinations in France around the introduction of a mandatory COVID certificate for several venues as compared to a reweighted Synthetic Control group Our World in Data,^26^ OxCGRT.^27^ Control pool: see methods.

We quantified the relative and absolute gains in vaccine uptake (Supplementary Table A5.1) of the anticipation of introduction (sum over 20 days before) and afterwards (sum over 40 days after). The anticipation effect in France meant that vaccine uptake exceeded the control country by 25,895 vaccines per million capita or in absolute terms, 1,749,589 doses. Vaccine uptake up to 40 days after certification exceeded the control country by 11,434 vaccines per million capita or 772,563 doses.

For Israel (Figure 3) we observe a similar shift before re-implementing the Green Pass. Three weeks before implementation, Israel reached the same level of vaccinations as the control group, providing evidence for an anticipation effect (although only towards average). The re-implementation coincided with a sharp increase in daily vaccinations directly afterwards and 40 days after implementation, rates were still more than 5,000 doses per million inhabitants higher. There was a limited anticipation effect, but a surge of 241,286 vaccines per million capita in total up to 40 days after which in absolute terms was 2,120,849 doses. We reflect on changes in eligibility by age groups in the next section.

**Figure 3.**
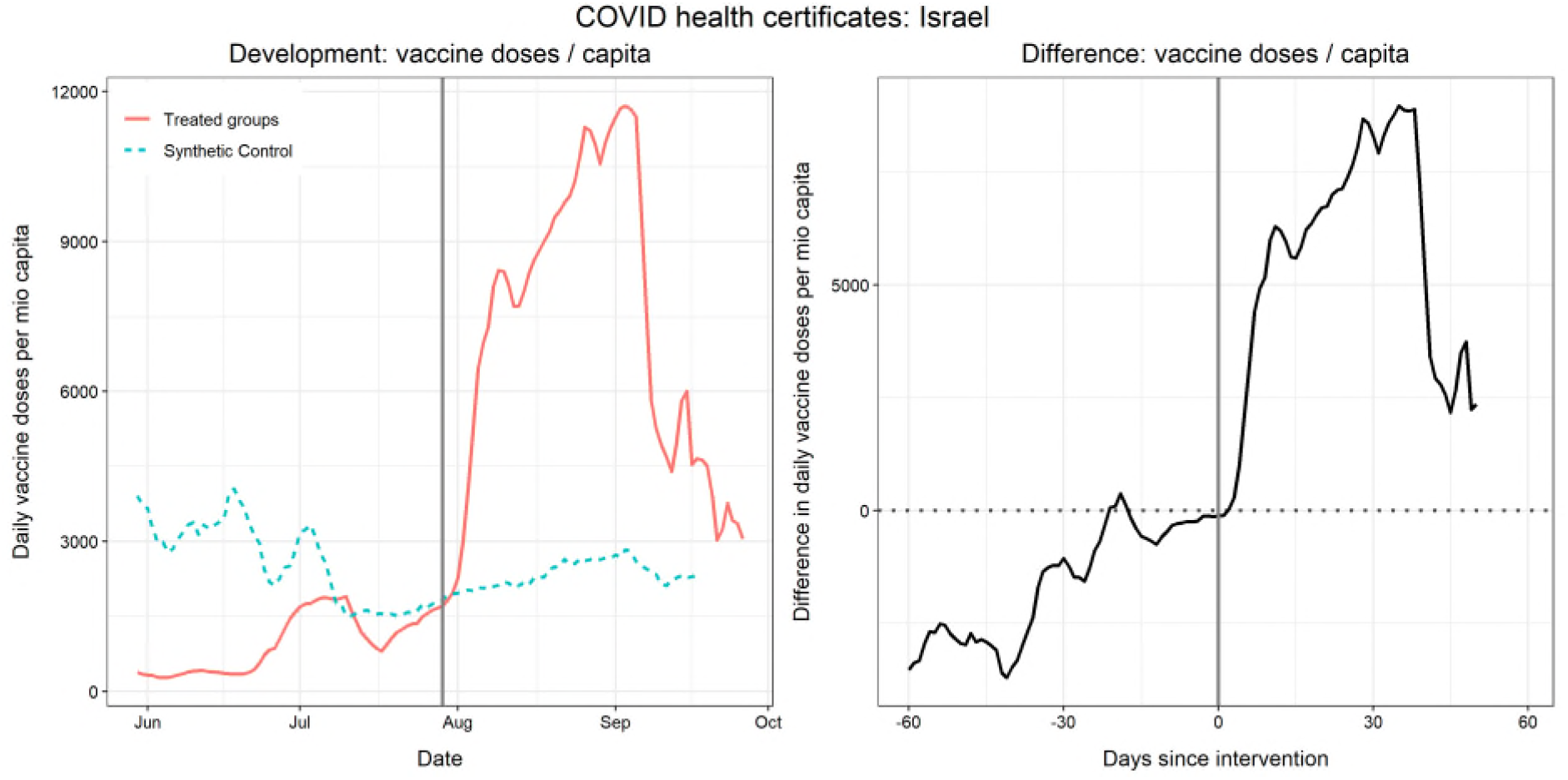
Daily new vaccinations in Israel around the introduction of a mandatory COVID certificate for several venues as compared to a reweighted Synthetic Control group Our World in Data,^26^ OxCGRT.^27^ Control pool: see methods.

For Italy (Figure 4), we again observe a shift from a below-average rate of daily vaccinations until the announcement of the Green Pass, followed by a decrease slightly below average of other countries after implementation. At 30 days after implementation, daily doses in Italy were around 1,500 doses above the synthetic control group, again confirming a positive relationship between certification and vaccine uptake.

**Figure 4.**
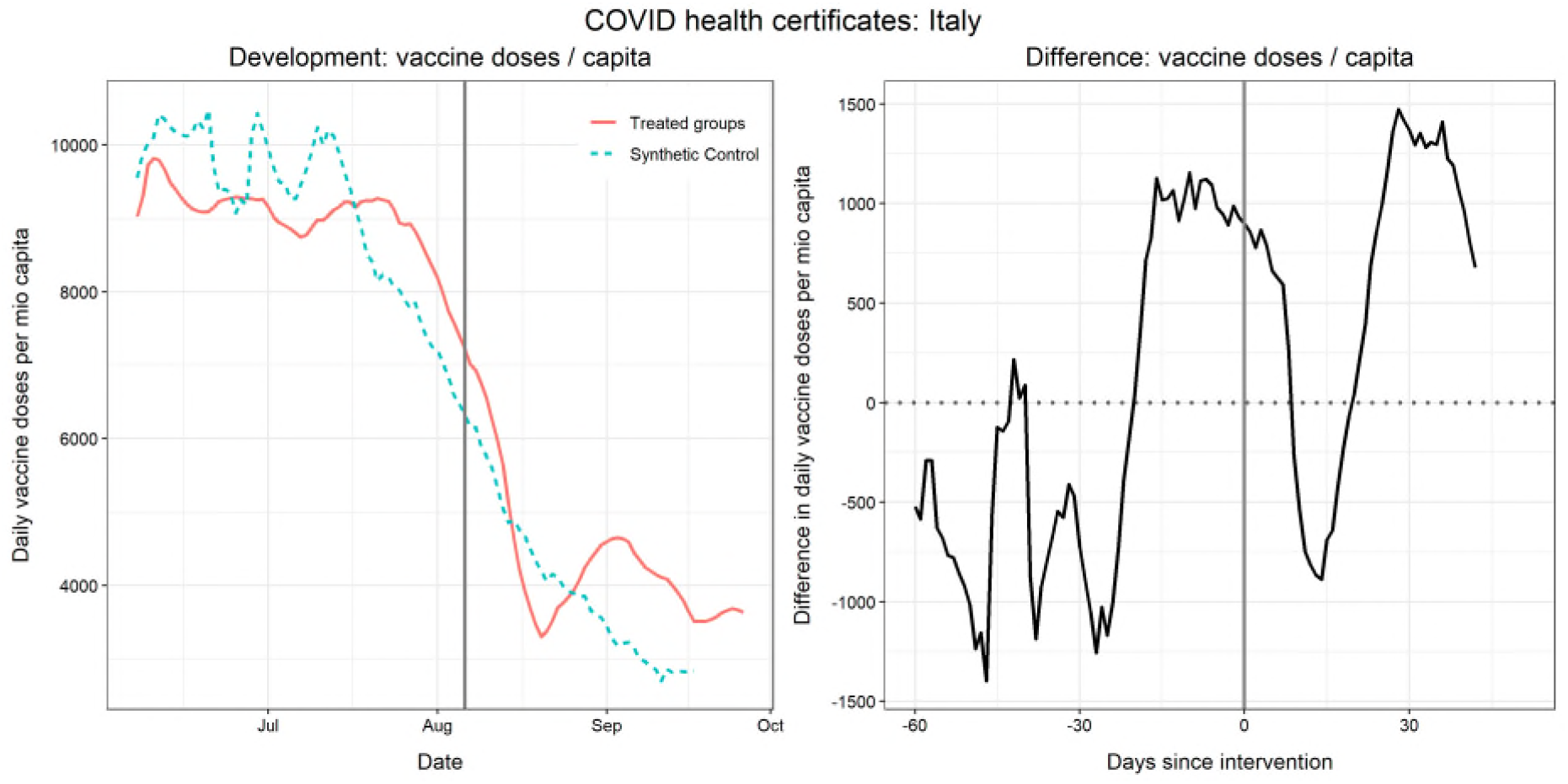
Daily new vaccinations in Italy around the introduction of a mandatory COVID certificate for several venues as compared to a reweighted Synthetic Control group. Data sources: Our World in Data,^26^ OxCGRT.^27^ Control pool: see methods.

Data in Switzerland (Figure 5) shows that around a month before the introduction of certificates, they had lower vaccination levels than control countries and daily doses approach an above-average trend briefly before the intervention. The observation period is too short for conclusions about longer-term trends, but does confirm an anticipation effect. Supplementary results (Figure A4.1) based on different estimation methods confirm the long lasting upward shift after certification.

**Figure 5.**
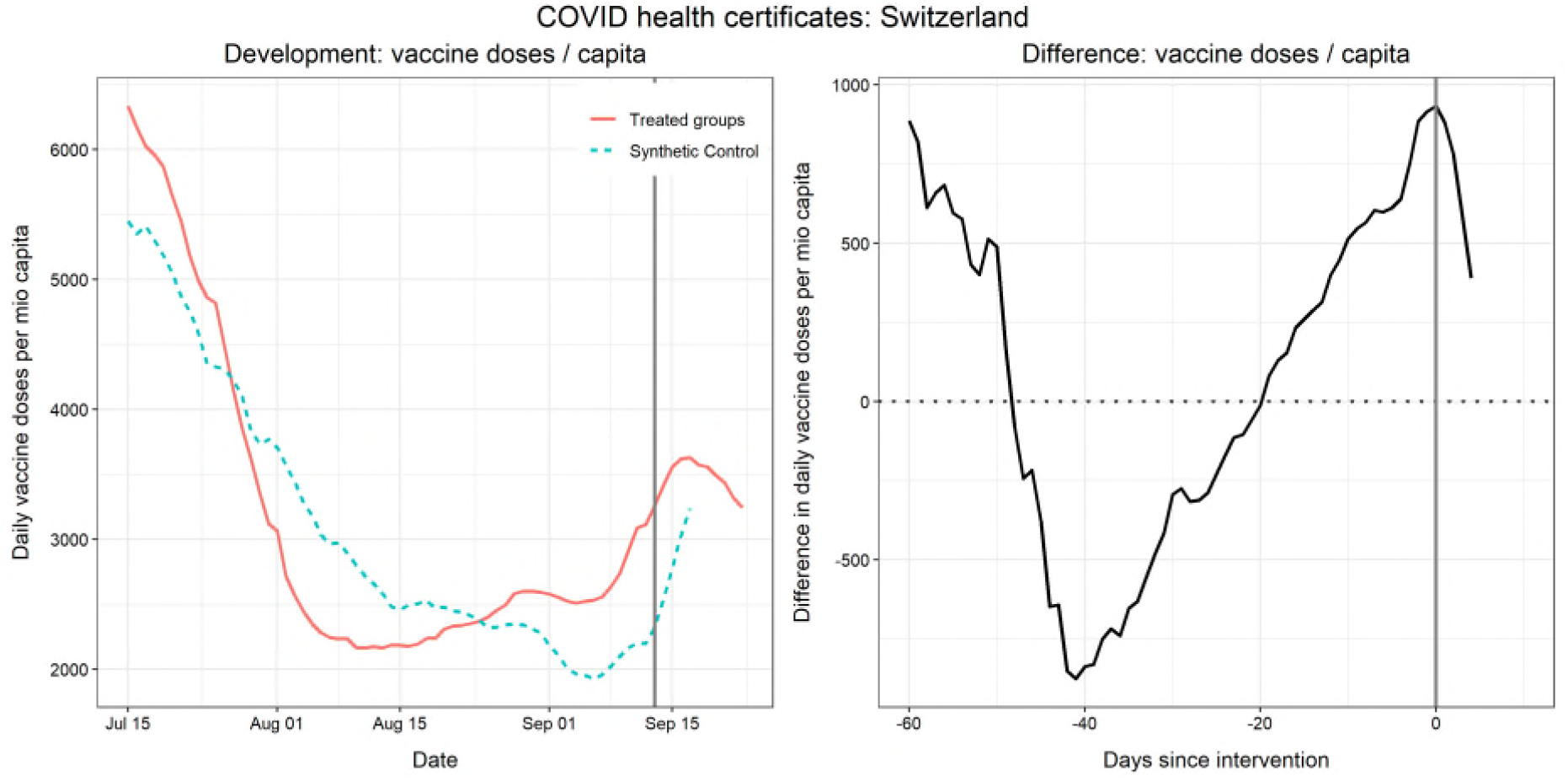
Daily new vaccinations in Switzerland around the introduction of a mandatory COVID certificate for several venues as compared to a reweighted Synthetic Control group Data sources: Our World in Data,^26^ OxCGRT.^27^ Control pool: see methods.

There are country-specific differences given different contexts and reasons for introducing certification, to increase testing (Denmark) or in response to rising cases (France, Germany, Belgium). In Denmark and Germany (Figures A1.1, A1.2), we do not observe an effect on vaccination rates.

Vaccination rates in Denmark were above others and the goal was to encourage regular testing, with the certificate now discontinued from 10 September given high vaccination levels. For Germany, vaccination rates increased three weeks after the intervention, but absolute differences remain small. Denmark introduced certificates very early in 2021 in times of limited supply. Germany has a federal structure of decision making, which resulted in location-specific rules and exemptions from certification based on local incidence rates (see Table A1). Germany was also in the midst of a federal election, where the two leading candidates publicly opposed certification.^5^ Both countries did not start from a below-average level of vaccine uptake compared to control countries before the intervention, indicating that the most leverage of certification was only if vaccination rates were below average than comparable countries.

We conducted secondary analyses on COVID-19 infections (Supplement A2) and concluded that the effect of certificates on reported cases is difficult to assess based on available data. For some countries (France, Germany, Italy) we observe a downwards trend after the intervention, whereas for others (Israel, Denmark) we observe a continued increase above the rate in other countries. The context of the pandemic trajectory when implementing the measure remains important (see Figure 1).

### Age-specific Analysis and Eligibility by age

Countries had different age-specific eligibility rollouts and first versus second doses (see Supplementary Table A2). Given the settings where certification was introduced such as nightclubs and large events, some age groups may respond differently. In Figure 6, we repeat previous analyses for France and Italy, but with age-specific vaccination rates. Both countries show a strong effect (anticipation and afterwards) for 25-49 year olds, while older age-groups seem less responsive, but also had a longer eligibility period to be vaccinated. In France, there is only a short period of increased uptake amongst the youngest ages after intervention. In Italy, the youngest age group (18-25) had an increase directly around the intervention and an even sharper increase 2-3 weeks after certification.

**Figure 6.**
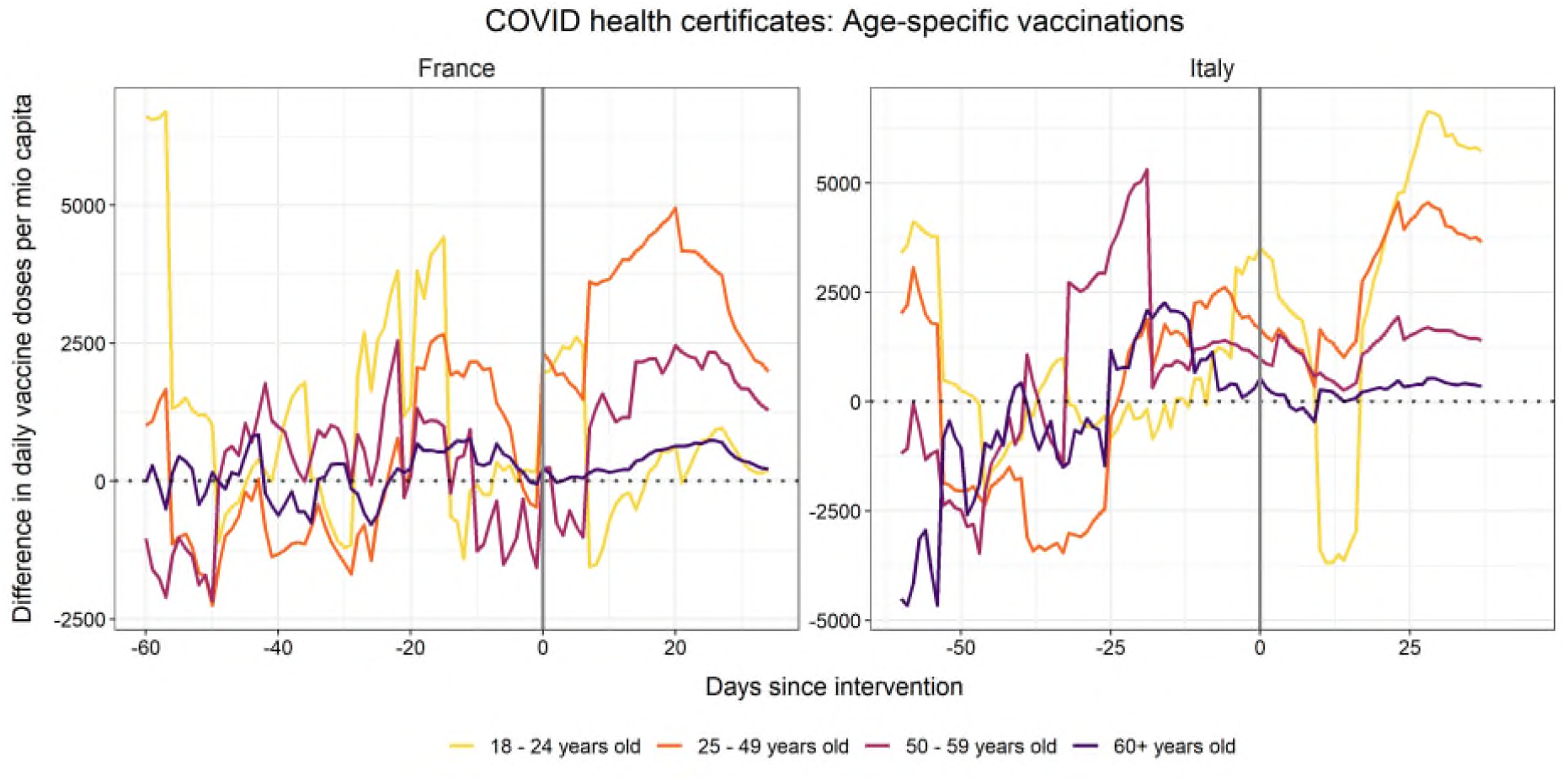
Daily new vaccinations by age group in France and Italy around the introduction of a mandatory COVID health certificate for several venues as compared to a reweighted Synthetic Control group Data sources: Our World in Data,^26^ OxCGRT,^27^ European Centre for Disease Prevention and Control.^14^ Control pool: see methods.

In addition to age-specific analyses based on synthetic control models, we also compiled age-specific data for France, Israel, Italy, and Switzerland (Figure 7), allowing a more detailed descriptive account of age groups. This fine-grained analysis confirms that those below 20 and 20-29 years old had increased uptake.

**Figure 7.**
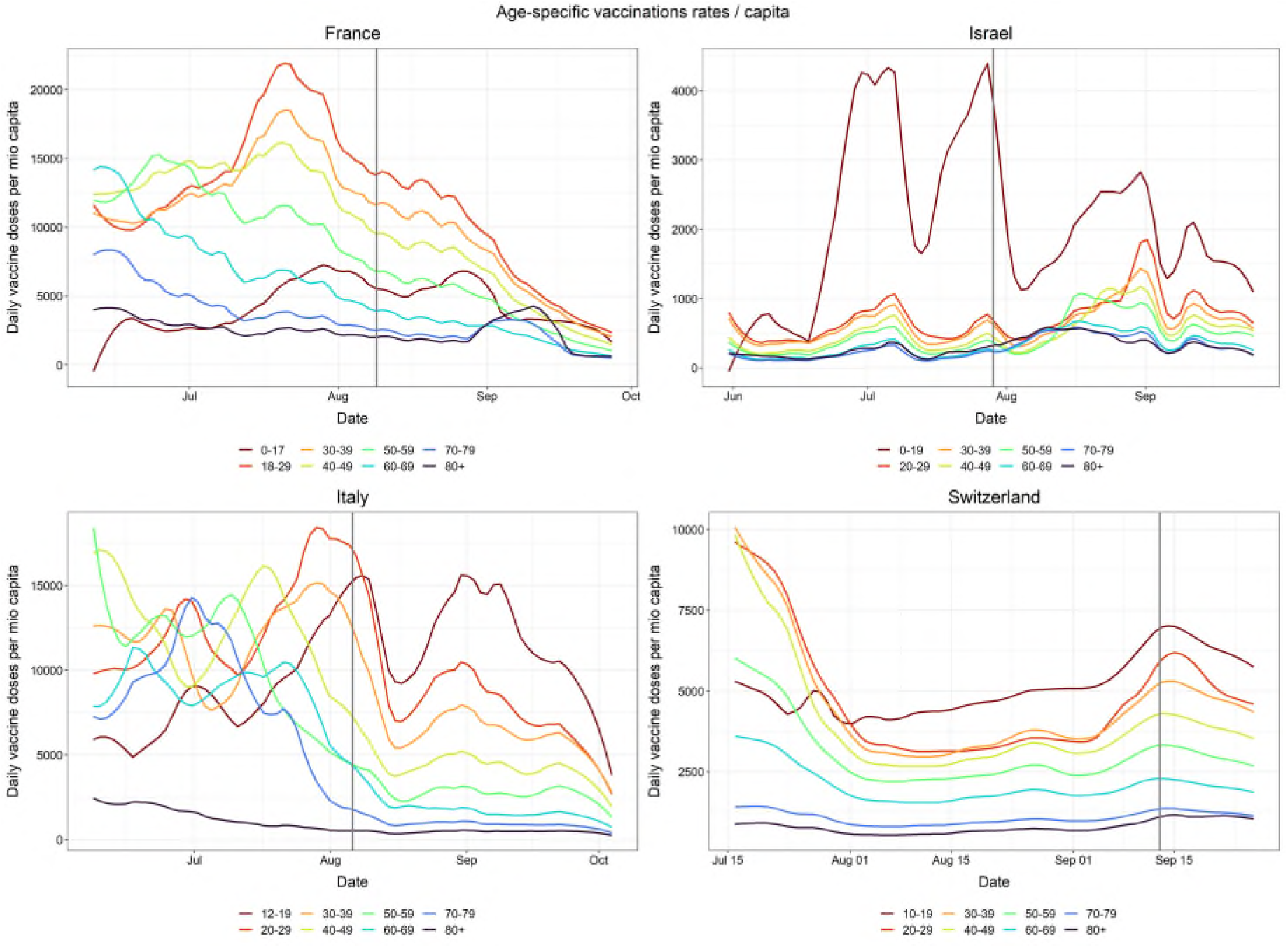
Descriptive development of daily new vaccinations by age group in France, Israel, Italy, and Switzerland around the introduction of a mandatory COVID certificate Data sources: Open platform for French public data,^15^ Government Database Israel,^16^ Extraordinary Commissioner for the Covid-19 emergency Italy,^17^ Federal Office of Public Health Switzerland.^18^ Note: the vertical lines represent the date of implementing mandatory COVID certificates.

One obvious concern is that our results are an artefact of age-related vaccine roll out and mirror the expanded eligibility to younger age groups during this period. To examine this, we link our findings to the timing of age-related eligibility (Supplementary Material Table A2). For France, children 12+ were eligible for vaccination on June 15, much earlier than the introduction of the certificate in August. In Italy, vaccines were available for children 12+ on August 16, which could partially explain the increase from mid-August, 2 weeks after certification was introduced. However, eligibility does not explain the pronounced increase amongst 20-29 year olds, eligible for a longer period. The two youngest age groups in Italy continued to have the highest vaccination rate amongst all age groups after the intervention. Findings hold for all doses, but also first dose only (Figure A3.1).

Although the sharp uptake amongst the youngest age groups in Israel could be attributable to second doses following the first spike, the intervention appears to be an uptake of first doses amongst teens below 20. Eligibility for vulnerable 5-11 year olds was introduced at the end of July, but this smaller group could not explain the sharp increase. Although children aged 12-15 were eligible on 06 June 2021, by the end of June only 2-4% of this age group were vaccinated. ^28^ After a surge in school infections, the Prime Minister urged citizens to vaccinate their children at the end of June, warning that the allotted doses would expire on July 9. Healthcare providers reported appointments for 12-15 year olds tripled directly, suggesting increased infections and a public announcement of urgency and waste may have prompted vaccinations.^28^ Supplementary analyses (Figure A3.1) show an increase around the introduction of certificates (July 29) for first doses only, which seems independent of the earlier announcement. Although smaller, we find an anticipation effect and a long-lasting increase in uptake after the intervention for first doses in Israel.

In Switzerland, vaccines were approved for 12-15 year olds on 04 June 2021, considerably before the introduction of stricter certification in mid-September. The actual rollout depended on the Canton, where for example, Zurich started to vaccinate younger ages on June 25 and Luzern only at the end of July. This increased rollout might have affected vaccine uptake of the first introduction of targeted certification rules (July 7), but it seems unlikely to affect our main intervention date (mid-September). It seems implausible that the staged introduction across Cantons is responsible for the small spike around the introduction of the first certification stage in early July.

In a secondary analysis, we use a natural experiment where Switzerland first introduced more targeted access restrictions (events > 1,000 participants and nightclubs), and later extended them to general situations and activities (Table A1). In Figure 8 the dashed line represents the first targeted introduction and the solid line to more general public settings (events >30 participants, entire hospitality sector, leisure activities). The targeted access restrictions appear to have only affected the youngest (below 20), while we barely observe a change amongst older groups. Extending these restrictions to more general activities continued to impact uptake in the youngest, but also uptake amongst older age groups (30-39, 40-49).

**Figure 8.**
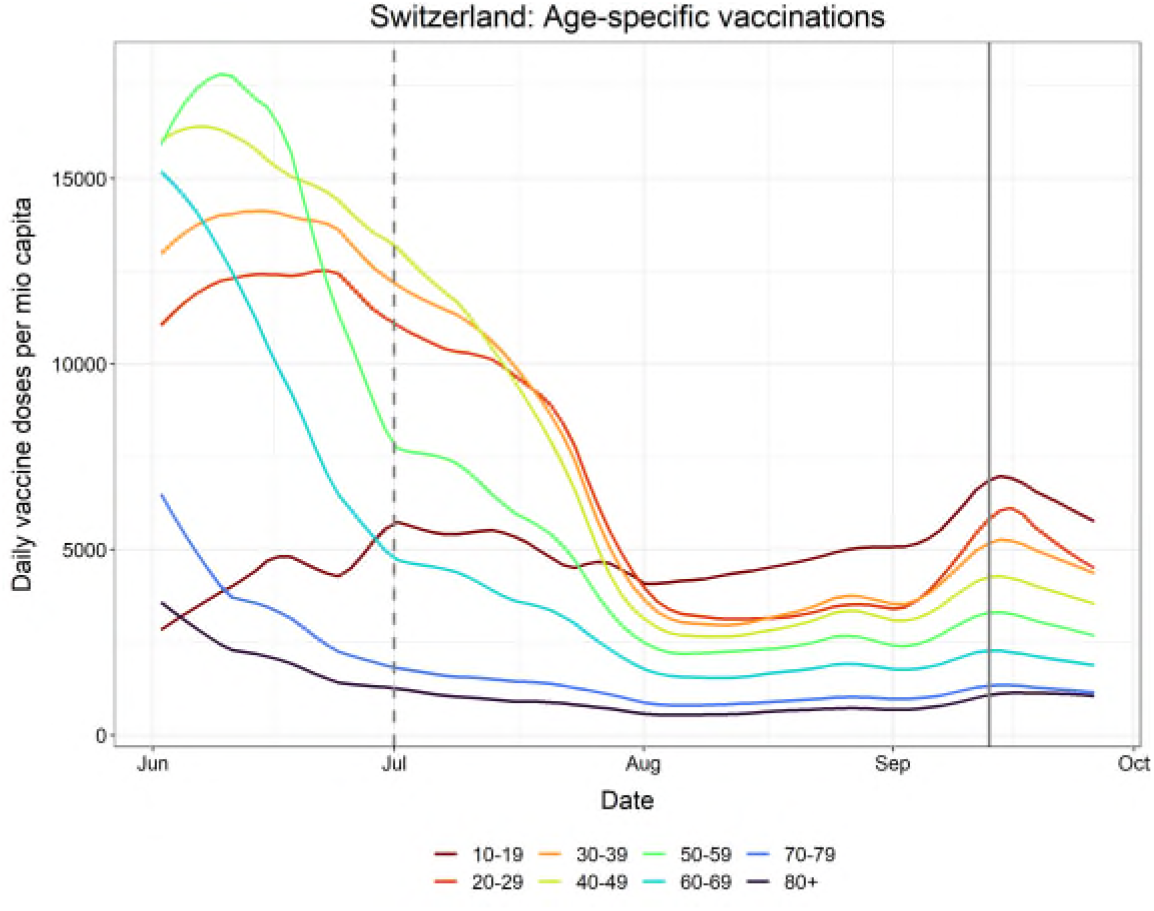
Descriptive development of daily new vaccinations by age group in Switzerland around the introduction of a targeted (dashed line) and general (solid line) mandatory COVID certificate Data sources: Federal Office of Public Health Switzerland.^18^ Note: vertical dashed line refers to t targeted access rules (events > 1,000 participants, nightclubs) and vertical solid line refers to introduction of general access restrictions (events > 30 participants, entire hospitality sector, and leisure activities). Source: Swiss Federal Department of Home Affairs.^29^

## Discussion

Mandatory COVID-19 certification led to a sharp increase in vaccination rates prior to implementation and had a long-lasting effect with above-average rates after implementation. The impact was dependent on the average pre-intervention levels of vaccine uptake in the country and reasons for introducing certification, with those that had lower average levels of uptake showing more pronounced effects (France, Italy, Israel), particularly in some age groups. There was no obvious effect in countries that had a higher uptake (Germany) or in times of limited supply or when it was introduced to increase testing (Denmark). The effect of certificates on COVID-19 infections is difficult to assess, but it is logical that encouraging people who attend higher risk venues to be vaccinated would reduce transmission risk.

The age effect is related to a combination of settings of where certificates were introduced that disproportionately impacted youth and changes in the eligibility criteria. A detailed age-specific analysis revealed that particularly those below the age of 20 had the highest increased uptake, followed by those 20-29 years, holding for both first and second doses. When introduced and incentives in particular settings (nightclubs, events >1,000 people), higher uptake was found mainly in the youngest age group in Switzerland and when certification was extended to broader settings (events >30 people, entire hospitality sector, leisure activities) and increase was also observed in older groups (30-49).

A recent survey in the UK concluded that vaccine passports were unlikely to shift vaccination intentions in younger age groups.^6^ Our results show the importance of moving away from self-reported intentions only to objective measures of behaviour. The current findings suggest that the introduction of COVID certification does increase vaccine uptake, particularly in younger age groups when linked to settings such as nightclubs and large events, and additionally in older age groups when applied to broader settings such as the entire hospitality sector. We also show that this intervention gains the most leverage if vaccination rates are below average. Our findings should be interpreted in relation to the national context of pre-existing levels of vaccine uptake, vaccine hesitancy, levels of trust in government and vaccinations, and pandemic trajectory. COVID certification is part of a constellation of multiple policy levers that could counter vaccine complacency and increase uptake.^8^ Youth and certain groups such as men and those from low socioeconomic environments demonstrate higher levels of vaccine complacency due to a lower assessment of the risk of COVID-19,^8,9^ suggesting certification could be one mechanism to increase uptake to reach vaccine immunity threshold levels to protect the broader population. Certification alone is not a silver bullet to increase vaccine uptake, with other measures such as geographically targeted interventions being more effective for certain groups.^30^

Limitations include a lack of access to granular daily age-based uptake for all countries and ability to examine confounders such as ethnicity or socioeconomic status. Certification was introduced at different phases during the pandemic for different reasons and across regional and national-level conditions with varying levels of age eligibility, supply, vaccine hesitancy, enforcement and variation in infections and mortality. France^31^ and Italy^32^ have a history of vaccine hesitancy;^31^ together with Israel incentivised vaccine uptake with the use of certification for desirable events or settings.^33^ Whereas Belgium’s aim was to avoid the reinstatement of restrictions.^34^ Transferability of results to other settings should account for contextual aspects listed above.

## Data Availability

Data sharing. All data is publicly available and listed in this document.

## Data sharing

All data is publicly available and listed in this document.

## Contributors

MCM and TR conceived the project and designed the study. MCM wrote the first draft of the manuscript, edited by both authors. TR conducted the statistical analyses. MCM and TR engaged in policy review. Both authors had full access to the data and final responsibility to submit for publication.

