## Supplementary Information for "The impact of mandatory COVID-19 certificates on vaccine uptake: Synthetic Control Modelling of Six Countries"

### 1. Summary of COVID-19 certification regulations, selected countries

**Table A1 Summary of COVID-19 certification announcements and regulations, selected countries (as of 29 Sept 2021)**

| Country | Announced | Introduced | Setting requirements | Exemptions, additional rules | Antigen testing | Selected sources |
|---|---|---|---|---|---|---|
| **Denmark** | 4 February 2021 | 6 April - 1 September 2021 (until 10 September for night clubs, large events) | 6 April: for access to hairdressers, beauty salons and driving schools,<br>21 April: outdoor hospitality,<br>6 May: indoor hospitality, cultural events,<br>21 May: all other venues | | Free of charge | End of covid pass:<br>https://www.euronews.com/2021/09/01/denmark-s-high-vaccination-rate-sees-covid-pass-ended-for-restaurants<br><br>Introduction of covid pass:<br>https://www.euronews.com/2021/04/06/covid-19-denmark-launches-coronapas-certificate-to-reopen-economy<br><br>Plans announced:<br>https://www.euronews.com/2021/02/04/covid-in-europe-denmark-announces-plans-for-a-corona-pass |
| **France** | 23 July 2021 | 09 August (or 29 July guidelines) depending on specific rules | Visiting hospitals, care homes, hospitality venues, events (>50 people), long-distance travel by plane, train or coach, campsites/hotels, malls,<br><br>Mandatory for workers who have contact with customers (since 30 August 2021), mandatory for workers in healthcare, fire service and police (since 15 September) | Local authorities able to put in place further restrictions on events | Free of charge for those covered by French social security | https://www.gouvernement.fr/en/coronavirus-covid-19<br><br>announcement:<br>https://www.lemonde.fr/politique/article/2021/07/23/passe-sanitaire-les-deputes-votent-la-mesure-controversee-nuit-d-apres-debats-a-l-assemblee_6089243_823448.html<br><br>mandatory for certain workers:<br>https://www.connexionfrance.com/French-news/French-health-pass-pass-sanitaire-to-become-obligatory-for-public-facing-employees-of-public-venues-from-August-30 |

| | | | | | | |
|---|---|---|---|---|---|---|
| | | | | | | August 09 reference: https://uk.ambafrance.org/Health-pass-and-vaccination-in-the-UK<br>29 July reference to other guidelines: https://www.gouvernement.fr/en/coronavirus-covid-19 |
| **Germany** | 3 August 2021 | 23 August 2021 | Visiting hospitals, care homes, hospitality venues, events, indoor parties and sports, for the use of body-related services (e.g., hairdresser, massages, etc.)<br>Vaccinated individuals do not need to self-isolate when entering Germany (even from high risk areas) | counties below a specific incidence rate exempted depending on federal state rules (7 day rate < 35 new infections per 100,000) | Free of charge until 11 October | https://www.bundesregierung.de/breg-de/aktuelles/bund-laender-beratung-corona-1949606<br><br>Speculation about prices of tests after 11 October: https://www.merkur.de/leben/gesundheit/corona-tests-kosten-ab-oktober-selbst-zahlen-pcr-antigentest-90978260.html<br><br>Announcement: https://www.tagesschau.de/inland/gesundheitsministerium-schutz-corona-101.html |
| **Israel** | 'old' green pass used previously reintroduced 29 July 2021<br><br>22 July 2021 (for reintroduction of 'old' green pass)<br><br>29 August 2021 (for introduction of 'new' green pass) | 19 February - 1 June 2021 initial 'old' green pass required<br><br>29 July 2021: Re-introduction of 'old' green pass<br><br>3 October 2021: 'new' green pass (valid for 6 months after | Cultural, sporting events, gyms, hospitality venues, conferences, tourist attractions, places of worship, events, higher education establishments | | Free of charge | https://www.bbc.co.uk/news/world-europe-56522408<br><br>https://lexatlas-c19.org/israel-is-the-green-pass-an-example-to-follow/ (initial green pass)<br><br>https://www.gov.il/en/departments/news/29072021-02<br><br>https://www.gov.il/en/departments/news/spoke_greenbadge220721 (announcement re-introduction)<br><br>New green pass (Ministry of health) https://corona.health.gov.il/en/directives/green-pass-info/<br><br>Announcement: |

| | | | | | | |
|---|---|---|---|---|---|---|
| | | vaccination or infection) | | | | https://www.gov.il/en/departments/news/29082021-01 |
| **Italy** | 22 July 2021 | 6 August 2021 | Train stations, culture/leisure venues, indoor sport, private parties, fairs, hospitality venues<br><br>Additionally:<br>Mandatory for all workers (from 15 October, announced 16 September) | | Free of charge | https://www.bbc.co.uk/news/world-europe-58590187<br><br>https://www.schengenvisainfo.com/news/italy-makes-covid-health-pass-mandatory-for-unvaccinated-people-including-foreign-tourists/<br><br>https://www.bbc.co.uk/news/world-europe-58590187<br><br>Testing:<br>https://www.thelocal.it/20210504/how-you-can-get-a-free-coronavirus-test-in-11-italian-cities/ |
| **Switzerland** | 19 May 2021<br><br><br><br>08 September 2021 | 7 July 2021<br><br><br><br>13 September 2021 | Obligatory:<br>Events > 1000 people, indoor events (e.g. clubs, discos, nightclubs)<br><br>Obligatory:<br>Events > 30 people indoor hospitality, culture/leisure venues (e.g., museums, swimming pools), private events in public venues (e.g., weddings) | Covid-certificate (valid for 1 year after vaccination/ 6 months after infection) | Free of charge until 10 October | https://www.newsd.admin.ch/newsd/message/attachments/68144.pdf<br><br>Announcement:<br>https://www.srf.ch/news/schweiz/leben-mit-dem-coronavirus-so-soll-das-covid-zertifikat-genutzt-werden<br>https://www.admin.ch/gov/de/start/dokumentation/medienmitteilungen.msg-id-85035.html<br><br>Testing:<br>https://www.bag.admin.ch/bag/de/home/krankheiten/ausbrueche-epidemien-pandemien/aktuelle-ausbrueche-epidemien/novel-cov/testen.html#-634686877 |

#### 2. Vaccine eligibility for children aged 12 and older by selected countries, 2021

**Table A2** Vaccine eligibility for children aged 12 and older by selected countries, 2021

| Country | Timing of age-related eligibility | Age related eligibility for younger groups | Exemptions, additional rules | Source |
|---|---|---|---|---|
| **Denmark** | mid July 2021 | 12-15 years | Earlier 16-17 year olds | https://www.euronews.com/next/2021/09/14/covid-vaccine-for-children-who-in-europe-is-leading-the-race https://www.sst.dk/en/English/Corona-eng/Vaccination-against-COVID-19/Who-should-be-vaccinated/12-15-year-olds https://www.thelocal.dk/20210715/children-aged-12-15-in-denmark-begin-covid-19-vaccination/ https://www.sst.dk/en/English/Corona-eng/Vaccination%20against%20COVID-19/Who%20should%20be%20vaccinated/16-18%20years |
| **France** | 15 June 2021 | 12 years and older | | https://www.euronews.com/2021/06/02/france-extends-covid-19-vaccination-to-12-18-year-olds https://www.euronews.com/next/2021/09/14/covid-vaccine-for-children-who-in-europe-is-leading-the-race |
| **Germany** | 07 June 2021 | 12 and older | Pfizer recommended only for children with certain chronic illnesses | https://www.reuters.com/world/europe/german-panel-gives-limited-approval-covid-19-shot-adolescents-2021-06-10/ https://www.reuters.com/business/healthcare-pharmaceuticals/eu-regulator-endorses-pfizer-biontech-covid-19-vaccine-adolescents-2021-05-28/ |
| | 16 August 2021 | 12 and older | No health restrictions | https://www.euronews.com/2021/08/02/germany-to-offer-covid-19-shots-for-all-kids-over-12 |
| **Israel** | 06 June 2021 | 12-15 year olds eligible | | https://www.jpost.com/israel-news/covid-did-israel-screw-up-not-vaccinating-children-ages-12-15-right-away-672203 |
| | 23 June 2021 | | After slow uptake Israeli PM urges citizens to vaccinate children, warning that allotted doses would expire on July 09 | https://www.reuters.com/world/middle-east/school-covid-19-cases-spur-israeli-parents-vaccinate-kids-2021-06-22/ |
| | 27 July 2021 | Vulnerable 5-11 year olds eligible | Vulnerable children only | https://www.timesofisrael.com/israel-to-start-vaccinating-kids-aged-5-11-who-have-severe-background-illnesses/ |
| | End August 2021 | plan to roll-out vaccines in schools | | https://www.thetimes.co.uk/article/children-in-israel-to-have-covid-vaccines-at-school-f565t8mjj https://www.reuters.com/world/middle-east/school-covid-19-cases-spur-israeli-parents-vaccinate-kids-2021-06-22/ |

| | | | | |
|---|---|---|---|---|
| **Italy** | 03 June 2021 | 12 years and older | Start of offering appointments to 12+ in some regions | https://www.thelocal.it/20210603/which-italian-regions-are-offering-covid-vaccine-appointments-to-all-from-thursday/ https://www.thelocal.it/20210811/covid-19-italy-to-vaccinate-12-18-year-olds-without-appointments/ |
| | Beginning July 2021 (Pfizer authorised late May 2021) | 12 years and older | Pfizer vaccines (authorised late May 12-15) began across most regions; Moderna (12-17) approved end July) | https://www.euronews.com/next/2021/09/14/covid-vaccine-for-children-who-in-europe-is-leading-the-race https://www.thelocal.it/20210603/which-italian-regions-are-offering-covid-vaccine-appointments-to-all-from-thursday/ |
| | 16 August 2021 | 12 years and older | Can get vaccine without an appointment | https://www.thelocal.it/20210811/covid-19-italy-to-vaccinate-12-18-year-olds-without-appointments/ |
| **Switzerland** | 04 June 2021 | Approved for 12-15 year olds | | https://www.bag.admin.ch/bag/de/home/krankheiten/ausbrueche-epidemien-pandemien/aktue[…]-cov/information-fuer-die-aerzteschaft/covid-19-impfung.html, https://www.swissmedic.ch/swissmedic/de/home/news/coronavirus-covid-19/covid-19-impfstoff-pfizer-biontech-fuer-jugendliche.html |
| | Late June, early July 2020 | 12 and older | Some areas started to offer to 12-15 such as canton of Zurich 25 June or Luzern end July | Zurich: https://www.zh.ch/de/gesundheit/coronavirus/coronavirus-impfung/impfgruppen.html#-1702582428 Luzern: https://gesundheit.lu.ch/themen/Humanmedizin/ Infektionskrankenheiten/Informationen_Coronavisu/Covid_Impfung |
| | 26 August 2021 | 12 and older | Recommended without restrictions | https://www.kinderaerzteschweiz.ch/Fuer-Mitglieder/Coronavirus---COVID-19 |

#### 3. Denmark and Germany

The results of the analyses for France, Israel, Italy and Switzerland were provided in the main body of the text. In Denmark, we observe a brief spike in vaccinations before the certificates were introduced. After introduction, daily vaccination rates were higher than among comparable countries. However, the daily variation is very high, which indicates that this effect might be driven by Denmark providing higher supplies than other countries since they are one of the earliest to implement vaccine passports.

Figure A1.1. Daily new vaccinations in Denmark around the introduction of a mandatory COVID health certificate for several venues as compared to a reweighted Synthetic Control group.

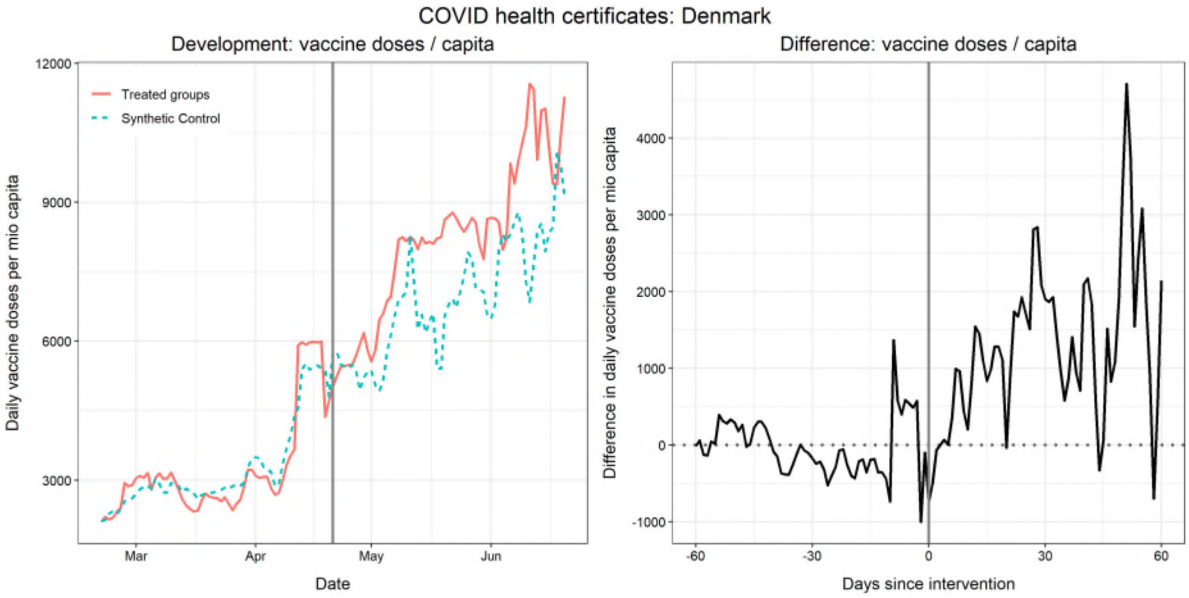

Data sources: Our World in Data (https://github.com/owid/covid-19-data), Oxford COVID-19 Government Response Tracker (https://github.com/OxCGRT/covid-policy-tracker). Control pool: AUT, BEL, CAN, CZE, ESP, FIN, GBR, GRC, HRV, IRL, LTU, LUX, NLD, NOR, POL, PRT, SVK, SVN, SWE, USA.

Germany provides an interesting exception for the patterns of daily vaccination doses before and after introducing mandatory health certificates. First, Germany did not introduce the certificates while following a below-average vaccination rate in comparison to other countries (note the small y range as compared to other figures/countries). Second, we see some slight spikes in vaccine uptake before introducing mandatory COVID certificates. Third, in contrast to remaining cases, daily vaccinations after the implementation fell below the rate observed in reweighted control countries. However, around 21 days after introduction, the vaccination uptake reaches above average levels (as the decline is smaller than the synthetic control unit. However, it is important to keep the local context in mind. First, Germany had different local measures in place already before the introduction of certificates. Second, federal states can exempt counties with an incidence rate below 35 from the restrictions related to the certificates. Third, the implementation was not linked to a lower-than-average vaccination rate prior to the intervention.

Figure A1.2. Daily new vaccinations in Germany around the introduction of a mandatory COVID health certificate for several venues as compared to a reweighted Synthetic Control group.

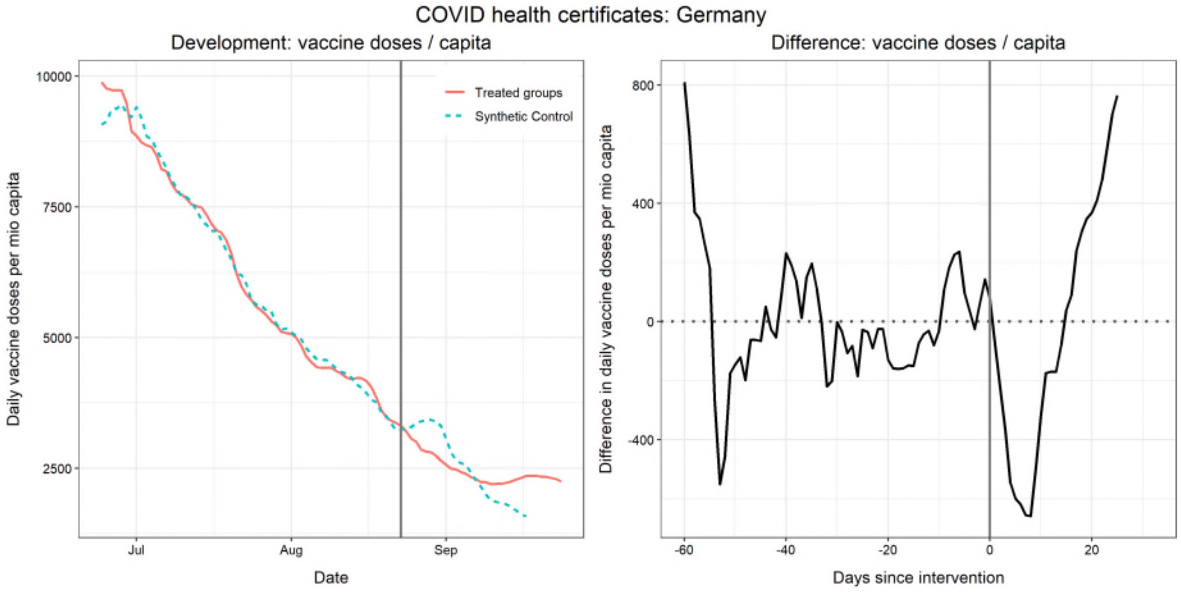

Data sources: Our World in Data (https://github.com/owid/covid-19-data), Oxford COVID-19 Government Response Tracker (https://github.com/OxCGRT/covid-policy-tracker). Control pool: AUT, BEL, CAN, CZE, ESP, FIN, GBR, GRC, HRV, IRL, LTU, LUX, NLD, NOR, POL, PRT, SVK, SVN, SWE, USA.

#### 4. Effect on COVID-19 cases

Figure A2.1. Daily COVID-19 cases in France around the introduction of a mandatory COVID health certificate for several venues as compared to a reweighted Synthetic Control group.

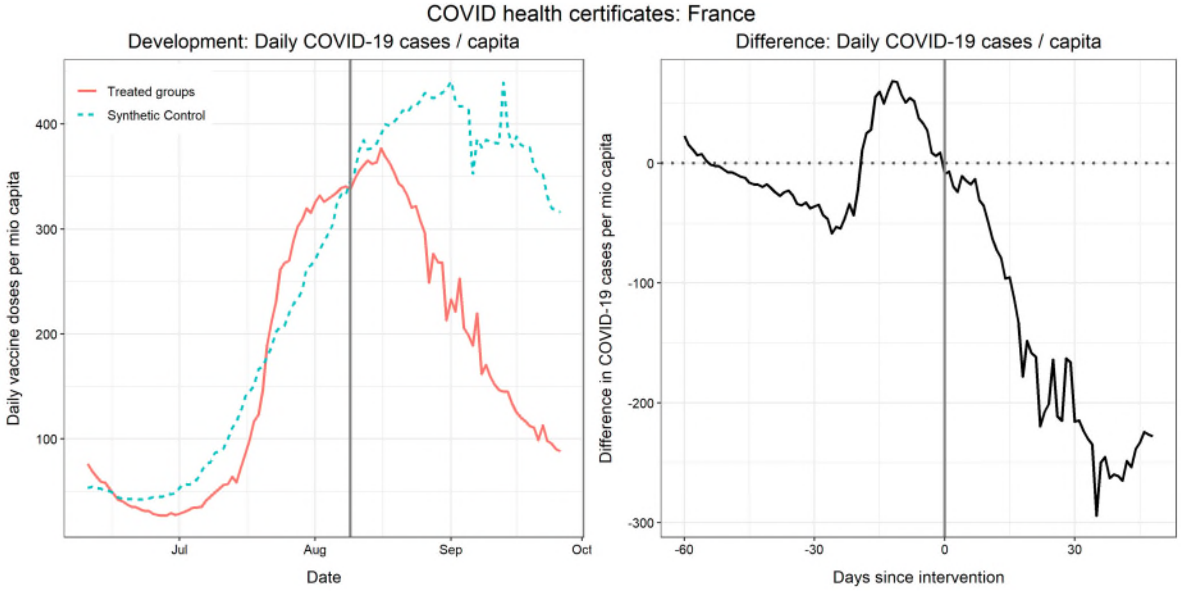

Data sources: Our World in Data ([https://github.com/owid/covid-19-data](https://github.com/owid/covid-19-data)), Oxford COVID-19 Government Response Tracker ([https://github.com/OxCGRT/covid-policy-tracker](https://github.com/OxCGRT/covid-policy-tracker)). Control pool: AUT, BEL, CAN, CZE, ESP, FIN, GBR, GRC, HRV, IRL, LTU, LUX, NLD, NOR, POL, PRT, SVK, SVN, SWE, USA.

Figure A2.2. Daily COVID-19 cases in Israel around the introduction of a mandatory COVID health certificate for several venues as compared to a reweighted Synthetic Control group.

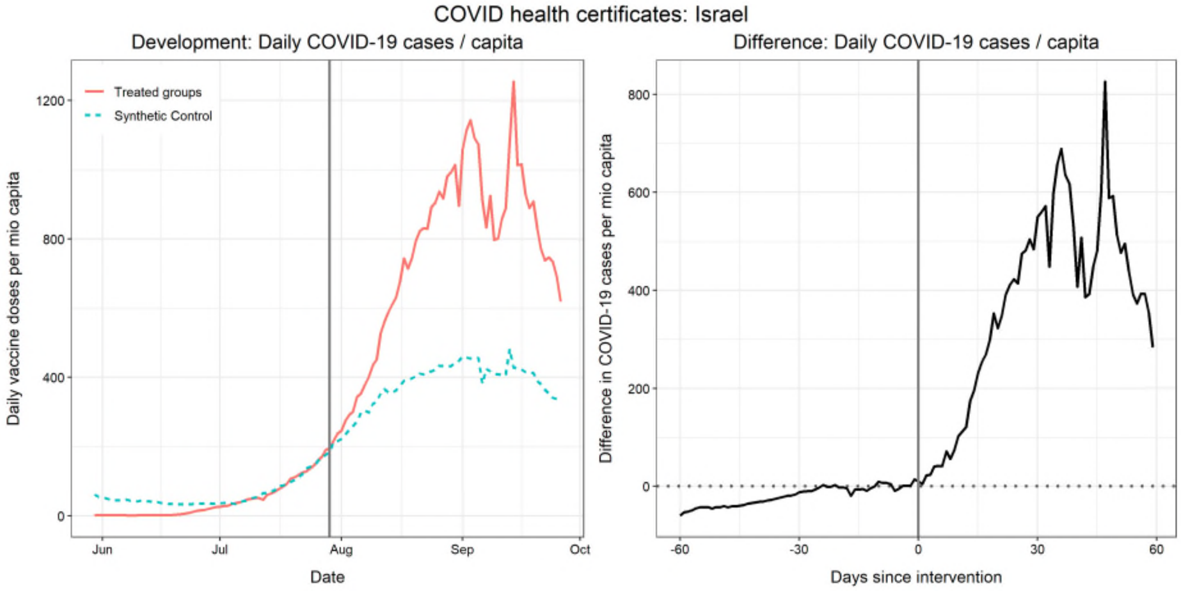

Data sources: Our World in Data ([https://github.com/owid/covid-19-data](https://github.com/owid/covid-19-data)), Oxford COVID-19 Government Response Tracker ([https://github.com/OxCGRT/covid-policy-tracker](https://github.com/OxCGRT/covid-policy-tracker)). Control pool: AUT, BEL, CAN, CZE, ESP, FIN, GBR, GRC, HRV, IRL, LTU, LUX, NLD, NOR, POL, PRT, SVK, SVN, SWE, USA.

Figure A2.5. Daily COVID-19 cases in Italy around the introduction of a mandatory COVID health certificate for several venues as compared to a reweighted Synthetic Control group.

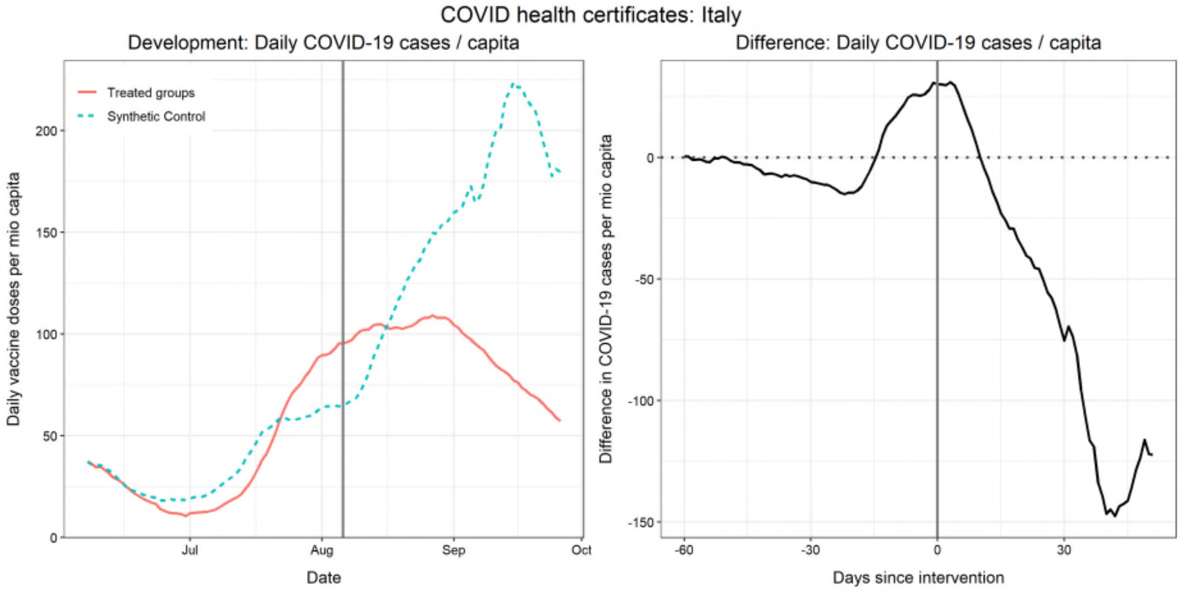

Data sources: Our World in Data (https://github.com/owid/covid-19-data), Oxford COVID-19 Government Response Tracker (https://github.com/OxCGRT/covid-policy-tracker). Control pool: AUT, BEL, CAN, CZE, ESP, FIN, GBR, GRC, HRV, IRL, LTU, LUX, NLD, NOR, POL, PRT, SVK, SVN, SWE, USA.

Figure A2.6. Daily COVID-19 cases in Switzerland around the introduction of a mandatory COVID health certificate for several venues as compared to a reweighted Synthetic Control group.

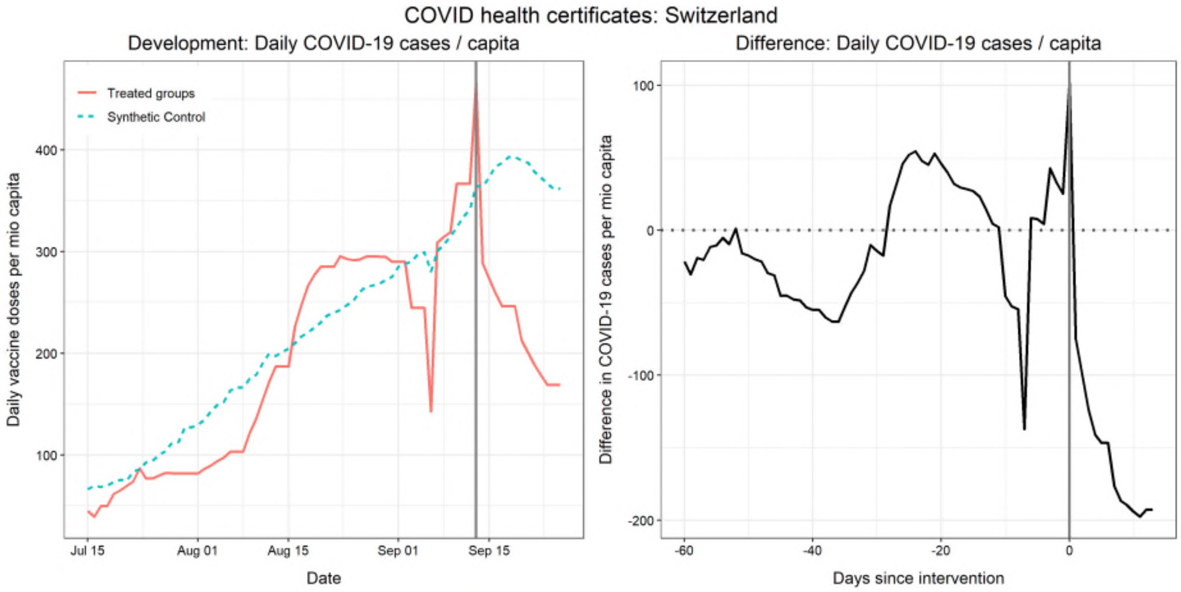

Data sources: Our World in Data (https://github.com/owid/covid-19-data), Oxford COVID-19 Government Response Tracker (https://github.com/OxCGRT/covid-policy-tracker). Control pool: AUT, BEL, CAN, CZE, ESP, FIN, GBR, GRC, HRV, IRL, LTU, LUX, NLD, NOR, POL, PRT, SVK, SVN, SWE, USA.

Figure A2.3. Daily COVID-19 cases in Denmark around the introduction of a mandatory COVID health certificate for several venues as compared to a reweighted Synthetic Control group.

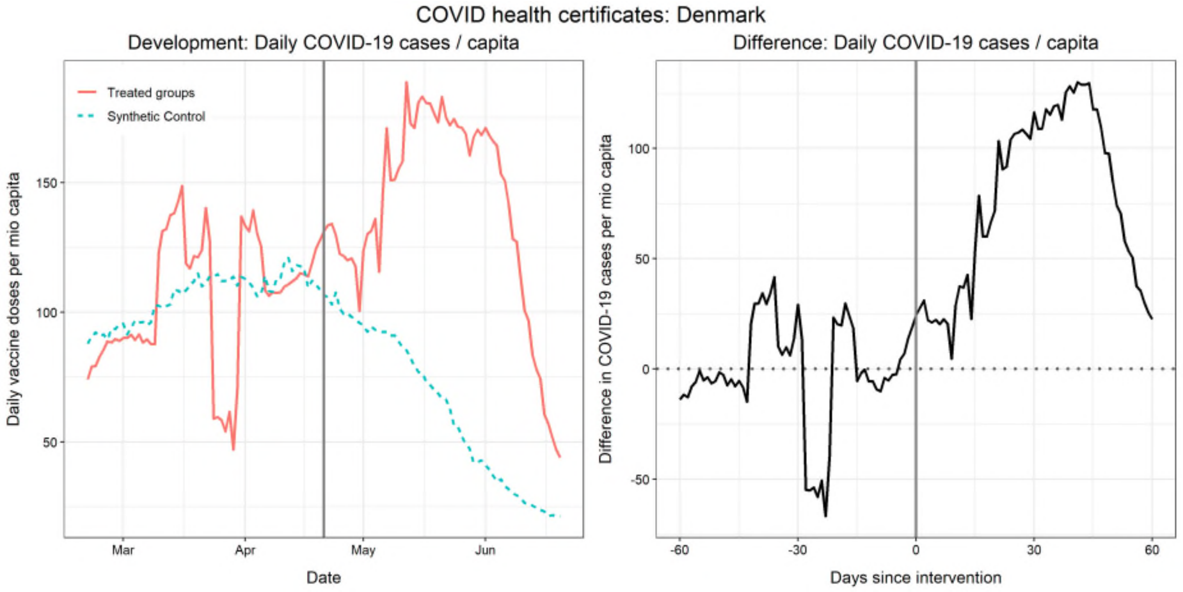

Data sources: Our World in Data (https://github.com/owid/covid-19-data), Oxford COVID-19 Government Response Tracker (https://github.com/OxCGRT/covid-policy-tracker). Control pool: AUT, BEL, CAN, CZE, ESP, FIN, GBR, GRC, HRV, IRL, LTU, LUX, NLD, NOR, POL, PRT, SVK, SVN, SWE, USA.

Figure A2.4. Daily COVID-19 cases in Germany around the introduction of a mandatory COVID health certificate for several venues as compared to a reweighted Synthetic Control group.

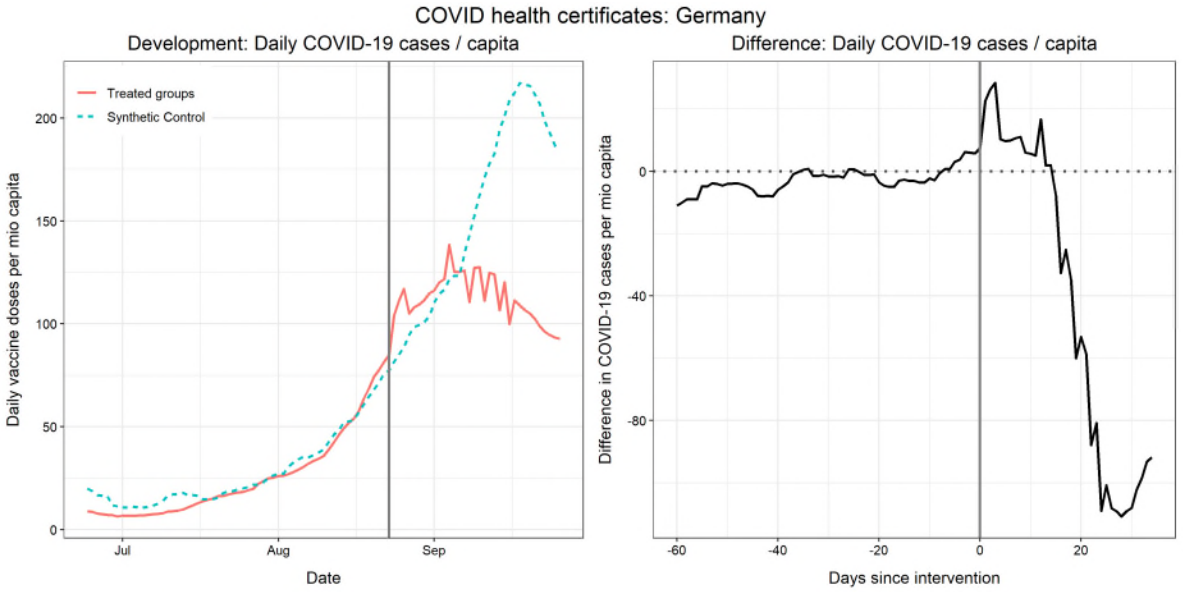

Data sources: Our World in Data (https://github.com/owid/covid-19-data), Oxford COVID-19 Government Response Tracker (https://github.com/OxCGRT/covid-policy-tracker). Control pool: AUT, BEL, CAN, CZE, ESP, FIN, GBR, GRC, HRV, IRL, LTU, LUX, NLD, NOR, POL, PRT, SVK, SVN, SWE, USA.

#### 5. Descriptive development of age-specific vaccinations

Figure A3.1. Descriptive development of daily new vaccinations by age group in France, Israel, Italy, and Switzerland for the first dose only around the introduction of a mandatory COVID health certificate.

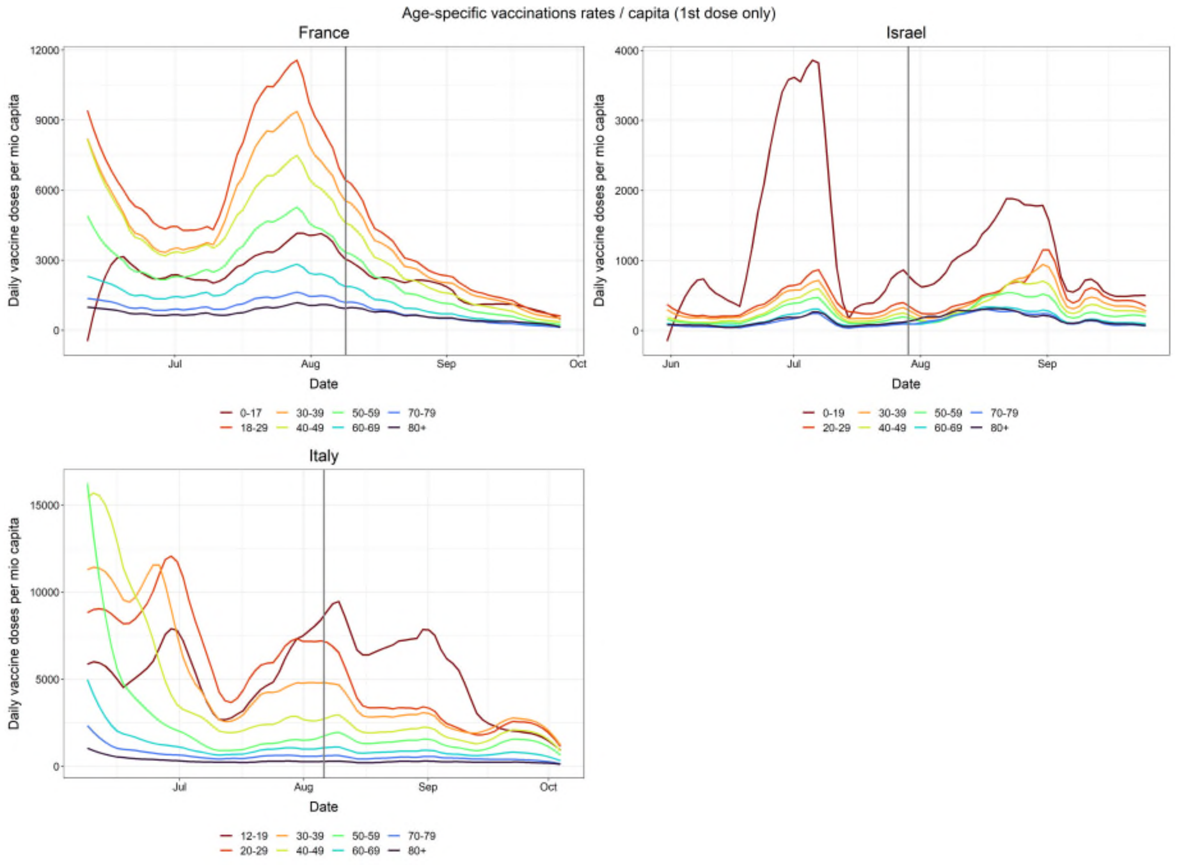

Data sources: Government Database Israel (https://data.gov.il/dataset/covid-19/resource/57410611-936c-49a6-ac3c-838171055b1f), Open platform for French public data (https://www.data.gouv.fr/en/datasets/donnees-relatives-aux-personnes-vaccinees-contre-la-covid-19-1/), Extraordinary Commissioner for the Covid-19 emergency Italy (https://github.com/italia/covid19-opendata-vaccini), Federal Office of Public Health Switzerland (https://www.covid19.admin.ch/en/vaccination/persons).

Note: the vertical lines represent the date of implementing mandatory COVID health certificates.

### 6. Generalized Synthetic Control

Figure A4.1. Generalized Synthetic Control method, comparing the case rates and vaccination rates in Germany, Denmark, France, and Israel to a counterfactual trajectory estimated based on all remaining countries.

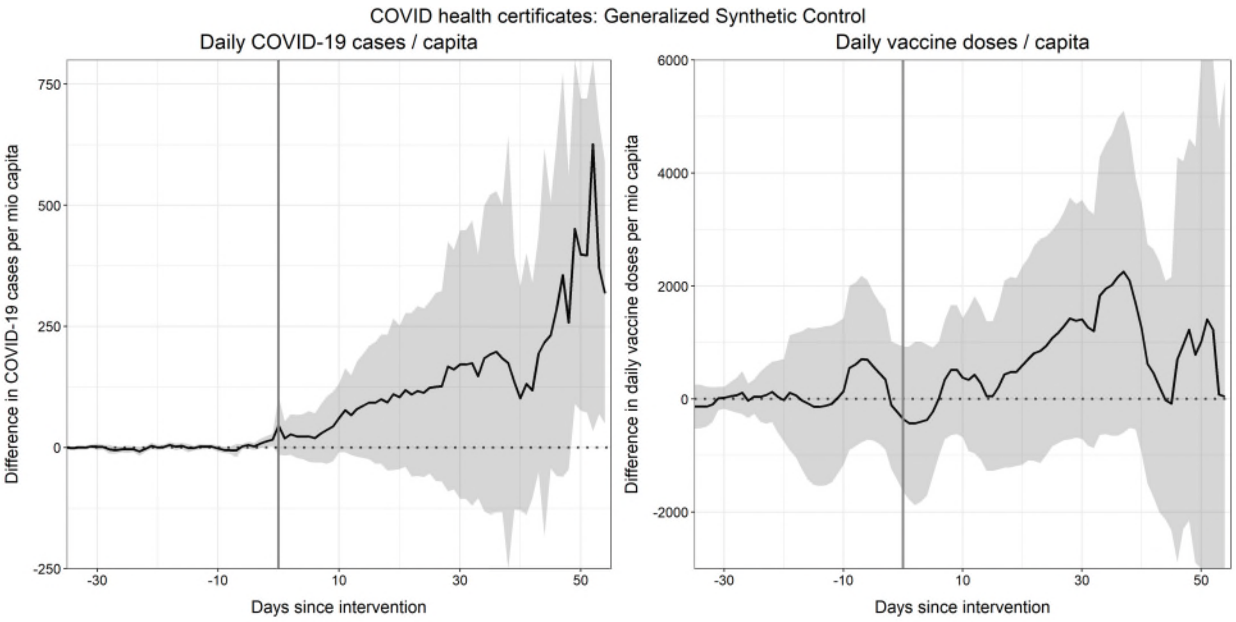

Data sources: Our World in Data (https://github.com/owid/covid-19-data), Oxford COVID-19 Government Response Tracker (https://github.com/OxCGRT/covid-policy-tracker). Treated: CHE, DEU, DNK, FRA, ITA, and ISR. Control: 163 remaining countries.

Note: Analysis of vaccinations controls for the 3-weeks lagged percentage of fully vaccinated. The treatment timing was preponed by three weeks to allow for potential anticipation effects. Shaded areas represent 95% confidence intervals based on a non-parametric bootstrap procedure (100 runs).

### 7. Summary measure of vaccine uptake

**Table A5.1.** Summary measure of the total difference in vaccinations between the synthetic control group and the treated country for an anticipation period and after the certificates were introduced.

| | Anticipation (sum over 20 days before) | | Uptake afterwards (sum over x days after) | | |
|---|---|---|---|---|---|
| | Total* | Per mio capita | Days | Total* | Per mio capita |
| France | 1,749,589 | 25,895 | 40 | 772,563 | 11,434 |
| Italy | 1,104,038 | 18,289 | 40 | 1,303,604 | 21,594 |
| Israel | -49,919 | -5,679 | 40 | 2,120,849 | 241,286 |
| Denmark | -2,248 | -387 | 40 | 243,203 | 41,836 |
| Germany** | -9,047 | -108 | 26 | -62,511 | -745 |
| Switzerland** | 77,687 | 8,914 | 5 | 31,166 | 3,576 |

Note: The sums show the daily differences between the synthetic control group and the respective treatment country (as shown in the right panels of Figures 2-5 of the main text). Bootstrapped standard errors are possible to compute for synthetic control methods but it is not possible given the low number of treated and control countries.

* Total describes the total number of administered vaccine doses as compared to the synthetic control group within the respective period. This highly depends on the total population, and the numbers are thus not comparable across the countries.

** For Germany and Switzerland we currently only observe a limited number of days after the intervention (26 and 5 days respectively). The uptake afterwards is thus not comparable to the remaining countries.
